# Induction of Cross-reactive Hemagglutination Inhibiting Antibody and Polyfunctional CD4+ T-cell Responses by a Recombinant Matrix-M-Adjuvanted Hemagglutinin Nanoparticle Influenza Vaccine

**DOI:** 10.1101/2020.05.11.20098574

**Authors:** Vivek Shinde, Rongman Cai, Joyce Plested, Iksung Cho, Jamie Fiske, Xuan Pham, Mingzhu Zhu, Shane Cloney-Clark, Nan Wang, Haixia Zhou, Bin Zhou, Nita Patel, Michael J Massare, Amy Fix, Michelle Spindler, David Nigel Thomas, Gale Smith, Louis Fries, Gregory M Glenn

## Abstract

**Background:** Recurrent reports of suboptimal influenza vaccine effectiveness have renewed calls to develop improved, broadly cross-protective influenza vaccines. Here, we evaluated the safety and immunogenicity of a novel, saponin (Matrix-M)-adjuvanted, recombinant hemagglutinin (HA) quadrivalent nanoparticle influenza vaccine (qNIV).

**Methods:** We conducted a randomized, observer-blind, comparator-controlled (trivalent high-dose inactivated influenza vaccine [IIV3-HD], or quadrivalent recombinant influenza vaccine [RIV4]), safety and immunogenicity trial of qNIV (in 5 different doses/formulations) in healthy adults aged ≥65 years. Vaccine immunogenicity was measured by hemagglutination-inhibition assays using reagents expressing wild-type HA sequences (wt-HAI) and cell-mediated immune (CMI) responses.

**Results:** A total of 1375 participants were randomized, immunized, and followed for safety and immunogenicity. Matrix-M-adjuvanted qNIV induced superior wt-HAI antibody responses against 5 of 6 homologous or drifted strains evaluated compared to unadjuvanted qNIV. Adjuvanted qNIV induced post-vaccination wt-HAI antibody responses at Day 28 that were: statistically higher than IIV3-HD against a panel of homologous or drifted A/H3N2 strains; similar to IIV3-HD against homologous A/H1N1 and B (Victoria) strains; and similar to RIV4 against all homologous and drifted strains evaluated. The qNIV formulation with 75 μg Matrix-M adjuvant induced substantially higher post-vaccination geometric mean fold-increases of influenza HA-specific polyfunctional CD4+ T-cells compared to IIV3-HD or RIV4. Overall, similar frequencies of solicited and unsolicited adverse events (AEs) were reported in all treatment groups.

**Conclusions:** qNIV with 75 μg Matrix-M adjuvant was well tolerated and induced robust antibody and cellular responses, notably against both homologous and drifted A/H3N2 viruses. Further investigation in a pivotal phase 3 trial is underway.

**SHORT SUMMARY:** We compared multiple formulations of a recombinant Matrix-M-adjuvanted nanoparticle influenza vaccine to 2 licensed influenza vaccines in older adults. The nanoparticle vaccine was well tolerated, and induced hemagglutination-inhibition antibody and CD4+ T-cell responses to vaccine-homologous and drifted A/H3N2 influenza viruses.

## INTRODUCTION

Seasonal influenza vaccination has been the cornerstone of prevention efforts to address the substantial health and economic burden of influenza [1,2]. However, recent developments, including several severe A(H3N2)-predominant influenza seasons; recurrent reports of poor field vaccine effectiveness from Europe, Canada, and the US; the increasingly recognized risk of antigenic mismatch arising from egg-based vaccine production; and the mounting challenge of predicting which viruses will circulate in the face of increasing strain diversity have undermined confidence in available influenza vaccines and reignited calls for developing improved, broadly cross-protective influenza vaccines [3-19]. These challenges have been acutely represented by contemporary circulating influenza A(H3N2) viruses because of their rapid rate of genetic and antigenic evolution, increased susceptibility to egg-adaptive mutations, and because they account for the majority of influenza-attributable morbidity and mortality [17,20-22], Additional challenges to overcome include potential modulatory effects of early-life immunological imprinting on vaccine effectiveness, limited durability of vaccine-induced protective immune responses, and limited induction of cellular immunity [4,7,23-25].

We recently described development of a novel saponin (Matrix-M)-adjuvanted recombinant hemagglutinin (HA) trivalent nanoparticle influenza vaccine (tNIV) produced in a Sf9 insect cell/recombinant baculovirus system that retains fidelity to wild-type (wt) circulating virus HA sequences, and contains conserved epitopes that stimulate broadly neutralizing antibodies (bnAbs) [26, 27], In a phase 1 study, tNIV demonstrated improved induction of wt-HAI antibody titers against A/H3N2 drift variants isolated over a 5-year period compared to an egg-derived, trivalent high-dose inactivated influenza vaccine (IIV3-HD) [28].

In the present phase 2 study, we further evaluated the safety and immunogenicity in adults ≥65 years of various doses and formulations of quadrivalent NIV (qNIV), with or without Matrix-M adjuvant, compared to 2 currently licensed influenza vaccines for which enhanced efficacy relative to standard IIV has been reported (IIV3-HD and quadrivalent recombinant HA influenza vaccine [RIV4]) [25].

## METHODS

### Study design

This randomized, observer-blind, comparator-controlled, dose and formulation optimization trial enrolled 1375 clinically stable adults ≥65 years across 14 US sites from September 24, 2018 to October 19, 2018. Eligible participants were randomized into 1 of 7 treatment groups, stratified by age (60 to <75 and ≥75 years), gender, and receipt of 2017-2018 seasonal influenza vaccine (Figure 1; Table S1). Inclusion and exclusion criteria are detailed in the Supplementary Appendix.

**Figure 1.**
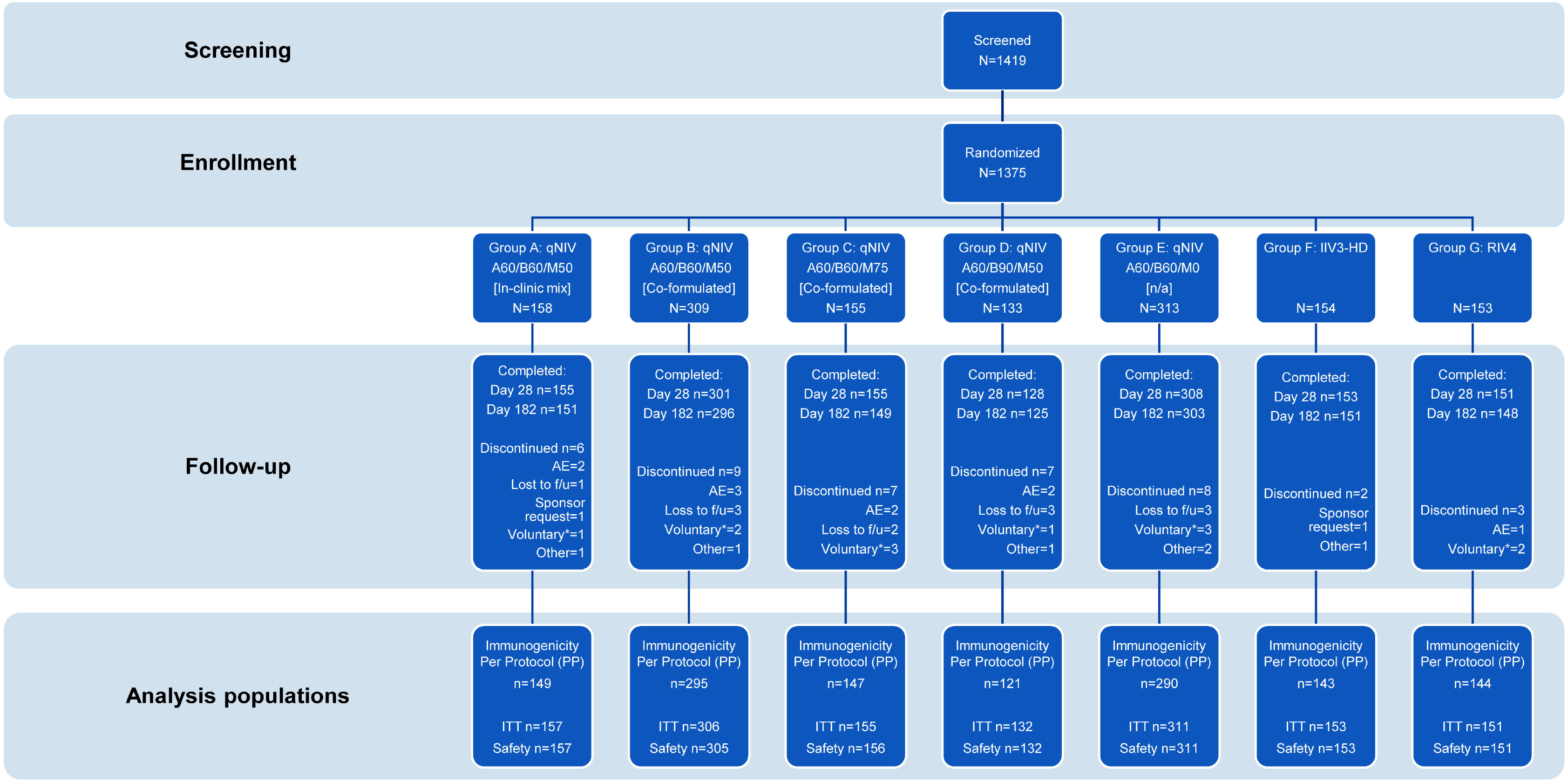
Flow diagram on screening, enrollment, and disposition of participants through the study. Abbreviations: A, influenza A strain HA antigen content in μg for each of A/H1N1 and A/H3N2 strains; AE, adverse event; B, influenza B strain hemagglutinin (HA) antigen content in μg for each of B/Victoria and B/Yamagata lineage strains; f/u, follow-up; IIV3-HD, trivalent high-dose inactivated influenza vaccine [Fluzone^®^ High-Dose]; ITT, intent-to-treat population; M, Matrix-M adjuvant content in μg; RIV4, quadrivalent recombinant influenza vaccine [Flublok^®^ Quadrivalent]; qNIV, quadrivalent recombinant nanoparticle influenza vaccine; Voluntary*, voluntary withdrawal unrelated to an AE. Definitions: safety population, defined as all participants who provided consent, were randomized, and received any investigational treatment, were used for all descriptive safety analyses; immunogenicity per protocol (iPP) population, defined as all participants in the safety population who received the assigned investigational treatment according to the protocol, had wt-HAI serology results for Days 0 and 28, and had no major protocol deviations affecting the primary immunogenicity outcomes as determined by the sponsor prior to database lock and unblinding, were used for all immunogenicity analyses; intent-to-treat (ITT) population, defined all participants in the safety population that provided any HAI serology data.

Investigational treatments comprised a single intramuscular injection of one of the following on Day 0 (Table 1): Group A: qNIV, with Matrix-M adjuvant and antigens mixed in the clinic just prior to administration; Group B: qNIV, pre-formulated with Matrix-M; Group C: qNIV, pre-formulated with high-dose Matrix-M; Group D: qNIV, pre-formulated with high-dose B antigens and Matrix-M; Group E: qNIV formulated without adjuvant; Group F: licensed IIV3-HD [trivalent Fluzone^®^ High-Dose]; or Group G: licensed RIV4 [FluBlok^®^ Quadrivalent] (Figure 1; Table S1). On Day 28, participants in Group E were administered a rescue injection with a licensed seasonal influenza vaccine; all other participants were administered a placebo injection on Day 28 to maintain trial blinding. Enrollment was divided into 3 stages to monitor safety (Supplementary Appendix).

**Table 1.** Summary of wt-HAI Antibody Responses by Treatment Group, Strain, and Time Point – Per Protocol Population.

| Treatment Group |  | A | B | C | D | E | F | G |
| --- | --- | --- | --- | --- | --- | --- | --- | --- |
| HA and Matrix Content |  | qNIV<br>A60/B60/M50 | qNIV<br>A60/B60/M50 | qNIV<br>A60/B60/M75 | qNIV<br>A60/B90/M50 | qNIV<br>A60/B60/M0 | IIV3-HD | RIV4 |
| Formulation |  | In-clinic mix | Co-formulated |  |  | NA | NA | NA |
| Subjects in Group (N) |  | N = 149 | N = 295 | N = 147 | N = 121 | N = 290 | N = 143 | N = 144 |
| Visit | Parameter |  |  |  |  |  |  |  |
| A/Michigan/45/2015 (H1N1) [Homologous Strain] |  |  |  |  |  |  |  |  |
| Day 0 | GMT | 50.1 | 47.9 | 55.3 | 48.7 | 48.7 | 50.5 | 44.0 |
|  | 95% CI | (44.6, 56.4) | (44.2, 51.9) | (49.0, 62.3) | (42.6, 55.8) | (44.8, 52.8) | (45.2, 56.4) | (39.5, 49.1) |
| Day 28 | GMT | 98.9 | 91.3 | 99.1 | 79.5 | 90.3 | 96.9 | 82.1 |
|  | 95% CI | (86.4, 113.1) | (82.5, 101.0) | (86.2, 114.0) | (68.6, 92.2) | (81.0, 100.5) | (84.5, 111.1) | (71.6, 94.2) |
|  | GMFR<sub>Post/Pre</sub> | 2.0 | 1.9 | 1.8 | 1.6 | 1.9 | 1.9 | 1.9 |
|  | 95% CI | (1.8, 2.2) | (1.8, 2.1) | (1.6, 2.0) | (1.5, 1.8) | (1.7, 2.1) | (1.7, 2.1) | (1.7, 2.1) |
| A/Singapore/INFIMH-16-0019/2016 (H3N2) [Homologous Strain] |  |  |  |  |  |  |  |  |
| Day 0 | GMT | 21.4 | 22.0 | 22.8 | 23.4 | 21.3 | 23.2 | 19.5 |
|  | 95% CI | (18.9, 24.3) | (20.0, 24.1) | (19.7, 26.3) | (20.2, 27.1) | (19.3, 23.5) | (20.4, 26.4) | (17.1, 22.2) |
| Day 28 | GMT | 65.8 | 65.4 | 64.2 | 59.4 | 50.8 | 46.5 | 66.6 |
|  | 95% CI | (55.2, 78.4) | (58.3, 73.5) | (54.2, 76.2) | (50.0, 70.5) | (45.0, 57.4) | (38.6, 55.9) | (54.9, 80.9) |
|  | GMFR<sub>Post/Pre</sub> | 3.1 | 3.0 | 2.8 | 2.5 | 2.4 | 2.0 | 3.4 |
|  | 95% CI | (2.6, 3.6) | (2.7, 3.3) | (2.4, 3.3) | (2.2, 3.0) | (2.1, 2.7) | (1.8, 2.3) | (2.8, 4.1) |
| B/Colorado/06/2017 (Victoria Lineage) [Homologous Strain] |  |  |  |  |  |  |  |  |
| Day 0 | GMT | 47.6 | 46.4 | 48.5 | 52.7 | 47.5 | 52.0 | 42.9 |
|  | 95% CI | (43.1, 52.6) | (42.8, 50.2) | (44.0, 53.6) | (46.7, 59.4) | (43.9, 51.3) | (46.1, 58.5) | (38.5, 47.7) |
| Day 28 | GMT | 86.8 | 83.2 | 89.6 | 95.5 | 73.2 | 93.2 | 83.3 |
|  | 95% CI | (77.1, 97.7) | (76.0, 91.0) | (79.3, 101.2) | (82.8, 110.3) | (67.6, 79.3) | (81.6, 106.5) | (73.2, 94.9) |
|  | GMFR<sub>Post/Pre</sub> | 1.8 | 1.8 | 1.8 | 1.8 | 1.5 | 1.8 | 1.9 |
|  | 95% CI | (1.6, 2.0) | (1.7, 1.9) | (1.7, 2.0) | (1.6, 2.1) | (1.4, 1.7) | (1.6, 2.0) | (1.7, 2.2) |
| B/Phuket/3073/2013 (Yamagata Lineage) [Homologous Strain] |  |  |  |  |  |  |  |  |
| Day 0 | GMT | 50.4 | 47.7 | 47.6 | 53.6 | 48.8 | 52.9 | 46.9 |
|  | 95% CI | (44.7, 56.7) | (44.4, 51.3) | (42.9, 52.9) | (46.9, 61.2) | (45.2, 52.7) | (47.3, 59.1) | (42.1, 52.2) |
| Day 28 | GMT | 108.5 | 101.7 | 104.9 | 113.8 | 87.5 | 64.5 | 102.0 |
|  | 95% CI | (96.2, 122.4) | (93.3, 110.8) | (92.4, 119.2) | (97.6, 132.6) | (79.5, 96.4) | (57.3, 72.6) | (88.6,117.4) |
|  | GMFR<sub>Post/Pre</sub> | 2.2 | 2.1 | 2.2 | 2.1 | 1.8 | 1.2 | 2.2 |
|  | 95% CI | (1.9, 2.4) | (2.0, 2.3) | (2.0, 2.5) | (1.8, 2.5) | (1.6, 2.0) | (1.1, 1.3) | (1.9, 2.5) |
| A/Switzerland/9715293/2013 (H3N2) [Drift Strain] |  |  |  |  |  |  |  |  |
| Day 0 | GMT | 54.6 | 60.0 | 60.9 | 60.6 | 58.6 | 62.3 | 54.8 |
|  | 95% CI | (47.8, 62.4) | (54.3, 66.2) | (52.7, 70.3) | (52.1, 70.5) | (52.8, 65.1) | (54.3, 71.6) | (47.8, 62.9) |
| Day 28 | GMT | 146.8 | 160.4 | 154.8 | 137.9 | 122.4 | 133.4 | 158.8 |
|  | 95% CI | (124.0, 173.9) | (143.9, 178.7) | (132.7, 180.7) | (117.2, 162.2) | (109.1, 137.3) | (111.2, 160.0) | (132.2, 190.9) |
|  | GMFR<sub>Post/Pre</sub> | 2.7 | 2.7 | 2.5 | 2.3 | 2.1 | 2.1 | 2.9 |
|  | 95% CI | (2.3, 3.1) | (2.4, 3.0) | (2.2, 2.9) | (2.0, 2.7) | (1.9, 2.3) | (1.9, 2.4) | (2.5, 3.4) |
| A/Wisconsin/19/2017 (H3N2) [Drift Strain] |  |  |  |  |  |  |  |  |
| Day 0 | GMT | 21.7 | 23.1 | 24.0 | 23.9 | 22.1 | 24.5 | 20.7 |
|  | 95% CI | (19.2, 24.6) | (21.1, 25.3) | (20.7, 27.7) | (20.9, 27.3) | (20.1, 24.4) | (21.6, 27.7) | (18.1, 23.6) |
| Day 28 | GMT | 61.1 | 63.2 | 63.0 | 58.2 | 50.1 | 46.1 | 64.3 |
|  | 95% CI | (51.8, 72.0) | (56.3, 70.9) | (53.5, 74.2) | (48.9, 69.2) | (44.4, 56.5) | (38.7, 55.1) | (53.5, 77.2) |
|  | GMFR<sub>Post/Pre</sub> | 2.8 | 2.7 | 2.6 | 2.4 | 2.3 | 1.9 | 3.1 |
|  | 95% CI | (2.4, 3.3) | (2.5, 3.0) | (2.3, 3.0) | (2.1, 2.9) | (2.0, 2.5) | (1.7, 2.1) | (2.6, 3.7) |

Abbreviations: CI, confidence interval; GMT, geometric mean titer; $GMR_{post/pre}$ , ratio of GMTs at point Day 28/Day 0; HA, hemagglutinin; N, number of subjects in per protocol population; n, number of subjects with non-missing hemagglutination- inhibition (HA) titer results at each visit; wt, wild-type. GMT was defined as the antilog of the mean of the log-transformed titer values for a given treatment group and time point. Individual antibody values recorded as below the lower limit of quantitation (LLOQ) were set to half LLOQ. $GMR_{post/pre}$ was defined as the ratio of 2 geometric mean titers within treatment group at 2 different time points between post-vaccination (Day 28) and pre-vaccination (Day 0). The 95% CI and P-value were obtained by paired t-test of $GMR = 1$ . Individual antibody values recorded as below the LLOQ were set to half LLOQ.

**Figure 2.**
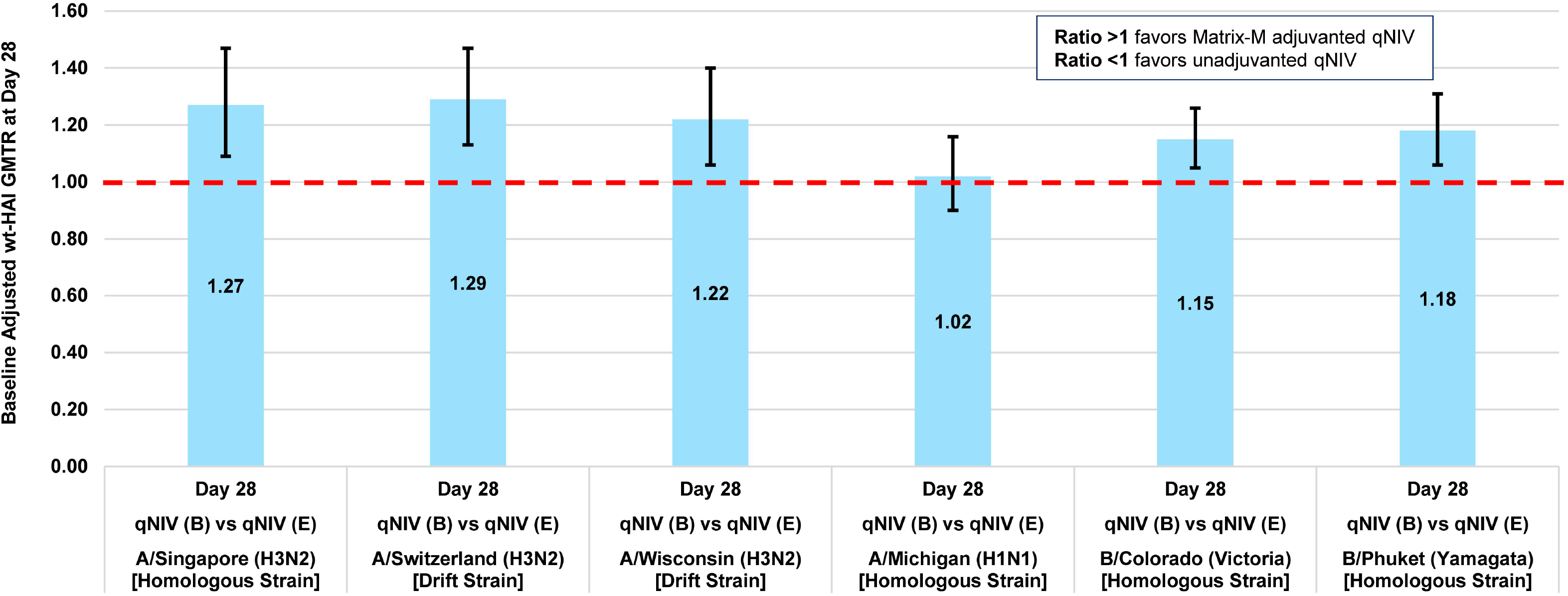
Demonstration of adjuvant effect – baseline adjusted ratio of Day 28 wt-HAI GMTs (GMTR) (Matrix-M-adjuvanted qNIV [Group B]/unadjuvanted qNIV [Group E]) Abbreviations: GMT, geometric mean titer; GMTR, ratio of GMTs; qNIV, quadrivalent recombinant nanoparticle influenza vaccine; wt-HAI, wild-type sequenced hemagglutinin inhibition antibody. Full strain names: A/Singapore/INFIMH-16-0019/2016 (H3N2); A/Switzerland/9715293/2013 (H3N2); A/Wisconsin/19/2017 (H3N2); A/Michigan/45/2015 (H1N1); B/Colorado/06/2017 (Victoria Lineage); B/Phuket/3073/2013 (Yamagata Lineage). Note: the primary immunogenicity objective of demonstrating an adjuvant effect required establishing immunogenic superiority of Group B (qNIV 60 μg HA x 4 strains with 50 μg Matrix-M1 adjuvant) relative to Group E (qNIV 60 μg HA x 4 strains without adjuvanted) by excluding values ≤1.0 at the lower 95% confidence bound for the baseline-adjusted ratio of Day 28 post-vaccination wt-HAI GMTs (GMTR; ie, GMT of Group B [adjuvant]/GMT of Group E [no adjuvant] at Day 28) for not less than 2 out of 6 influenza strains (ie, any 2 of 4 vaccine-homologous strains and/or 2 antigenically drifted influenza strains), while no other strain(s) demonstrated GMTRs which were significantly <1.0.

Participants had scheduled follow-up during up to 7 in-person clinic visits or telephone calls spanning 182 days to measure vital signs, perform physical exams, report adverse events (AEs), record concomitant medication changes, and collect blood samples for immunogenicity analyses (Table S2).

### Ethics and oversight

All participants provided written informed consent. The trial was reviewed and approved by Quorum Review, Seattle, WA.

### Study objectives

The primary objectives were to describe the safety and tolerability of each test vaccine, and to demonstrate a Matrix-M adjuvant effect by demonstrating the immunogenic superiority of qNIV (60 μg HA per A and B strain) formulated with 50 μg Matrix-M1 per dose compared to qNIV (60 μg HA per A and B strain) *without* adjuvant, based on Day 28 post-vaccination wt-HAI antibody responses against 4 vaccine-homologous (2 influenza A and 2 influenza B strains) and/or 2 antigenically drifted A/H3N2 strains. Additional objectives included describing post-vaccination wt-HAI antibody response (secondary objective) and cellular immune response (exploratory objective) of qNIV relative to IIV3-HD and RIV4 at various time points (Supplementary Appendix).

### Vaccines

Details of qNIV formulations, Matrix-M adjuvant, and comparators (IIV3-HD and RIV4) are provided in the Supplementary Appendix.

### Immunogenicity assessments

Blood samples were collected from participants on Days 0, 28, 56, and 182 for serological analyses. To measure the most biologically relevant vaccine-induced HAI antibody responses -- those against circulating wt hemagglutinins without egg-adapative antigenic changes -- we previously developed the wt-HAI assay as a modification of the classical HAI method by utilizing recombinant wt HA virus-like particles as the agglutinating agent (Supplementary Appendix) [28].

Peripheral blood mononuclear cells (PBMCs) were collected in a subset of 189 participants (comprising participants from 3 study sites [^~^63 per site]) at Days 0 and 7 for cell-mediated immunity (CMI) analyses. Due to limited availability of PBMCs per participant, only a subset of informative treatment groups and strains were tested for CMI based on the results of the wt-HAI data (Supplementary Appendix).

### Safety assessments

Safety follow-up consisted of collection of all solicited local and systemic AEs over 7 days post-Day 0 dosing; all AEs through 28 days post-Day 0 dosing; and all medically attended events (MAEs), serious AEs (SAEs), and significant new medical conditions (SNMCs; including immunologically mediated AEs of special interest) through Day 182 post-Day 0 dosing.

### Statistical analysis

Safety, immunogenicity per protocol (iPP), and intent-to-treat (ITT) populations are described in Figure 1 and the Supplementary Appendix.

Data concerning wt-HAI titers were expressed as geometric mean titers (GMTs), geometric mean fold-rise (GMFRp_ost/pre_), between-group ratio of GMTs (GMTR), seroconversion rate (SCR), and seroprotection rate (SPR) (Supplementary Appendix).

For CMI responses, measured by intracellular cytokine staining (ICCS), peripheral blood CD4+T-cell producing interleukin-2 (IL-2), interferon gamma (IFN-γ), and/or tumor necrosis factor alpha (TNF-α) cytokines following in vitro re-stimulation with influenza vaccine-homologous (A/Singapore/FIMH-16-0019/2016[H3N2]; A/Michigan/45/2015[H1N1]; B/Colorado/06/2017[Victoria]), or drifted (A/Wisconsin/19/2017[H3N2]) strain-specific HAs were reported as median cell counts, geometric mean cell counts (GMCs), and GMFR_Post/pre_ of double-cytokine (2 of 3: IL-2, IFN-γ, or TNF-α) or triple-cytokine producing (IL-2, IFN-γ, and TNF-α) influenza strain-specific CD4+ T-cells (for each individual strain). Between-group differences were reported as the ratio of GMCs (GMCR) at Day 7 of double- or triple-cytokine responses (and associated 90% CIs) [29].

The success criterion for the primary immunogenicity objective is described in Figure 2 and the Supplementary Appendix. A total sample size of 1350 was selected to provide ≥80% power to achieve the primary immunogenicity objective of demonstrating an adjuvant effect.

All statistical analyses were performed in SAS statistical software (Version 9.4).

## RESULTS

### Study participants

A total of 1375 participants were enrolled and randomized into 1 of 7 treatment groups (Figure 1; Table S1). Twelve (<1%) participants discontinued from the trial through Day 28 (Figure 1). Baseline characteristics of participants were similar across treatment groups. Mean age ranged from 71.8–72.9 years (Table S3). The proportion of females in each treatment group varied between 49% and 65%. The majority of participants in each group were white and had received influenza vaccine during the previous season (85–89%).

### Safety

The safety and reactogenicity profile was comparable between treatments groups through Day 182 (Table 2). All treatments were well tolerated. Rates of all solicited AEs were comparable (27.3–38.9%) across treatment groups; severe solicited local or systemic AEs were infrequent (<3.5% in any group). The most common solicited local AEs were injection site pain (10.3–19.3%), swelling (3.8–10.2%), and redness (2.6–7.6%); common solicited systemic AEs included headache (7.4–14.7%), muscle pain (4.6–13.1%), and fatigue (5.1–10.8%) (Table S6). MAEs were reported in a similar proportion of participants across all groups (24–32% qNIV; 22% IIV3-HD; 27% RIV4), with no apparent clustering by diagnosis or treatment group. Overall, SAEs rates were as expected given the age of the enrolled population, and reported in <10% of participants across the entire study. SAEs occurred in 5.1–9.1% of participants in the adjuvanted qNIV groups (pooled rate 5.9%), compared to 3.9% of IIV3-HD participants and 2.0% of RIV4 participants (Table 2; Table S7). No SAEs were considered related to study treatment in any treatment group, among a total of 63 SAEs reported in 59 participants. Out of 59 participants, 7 died with all events assessed as not related by the study investigators.

**Table 2.**
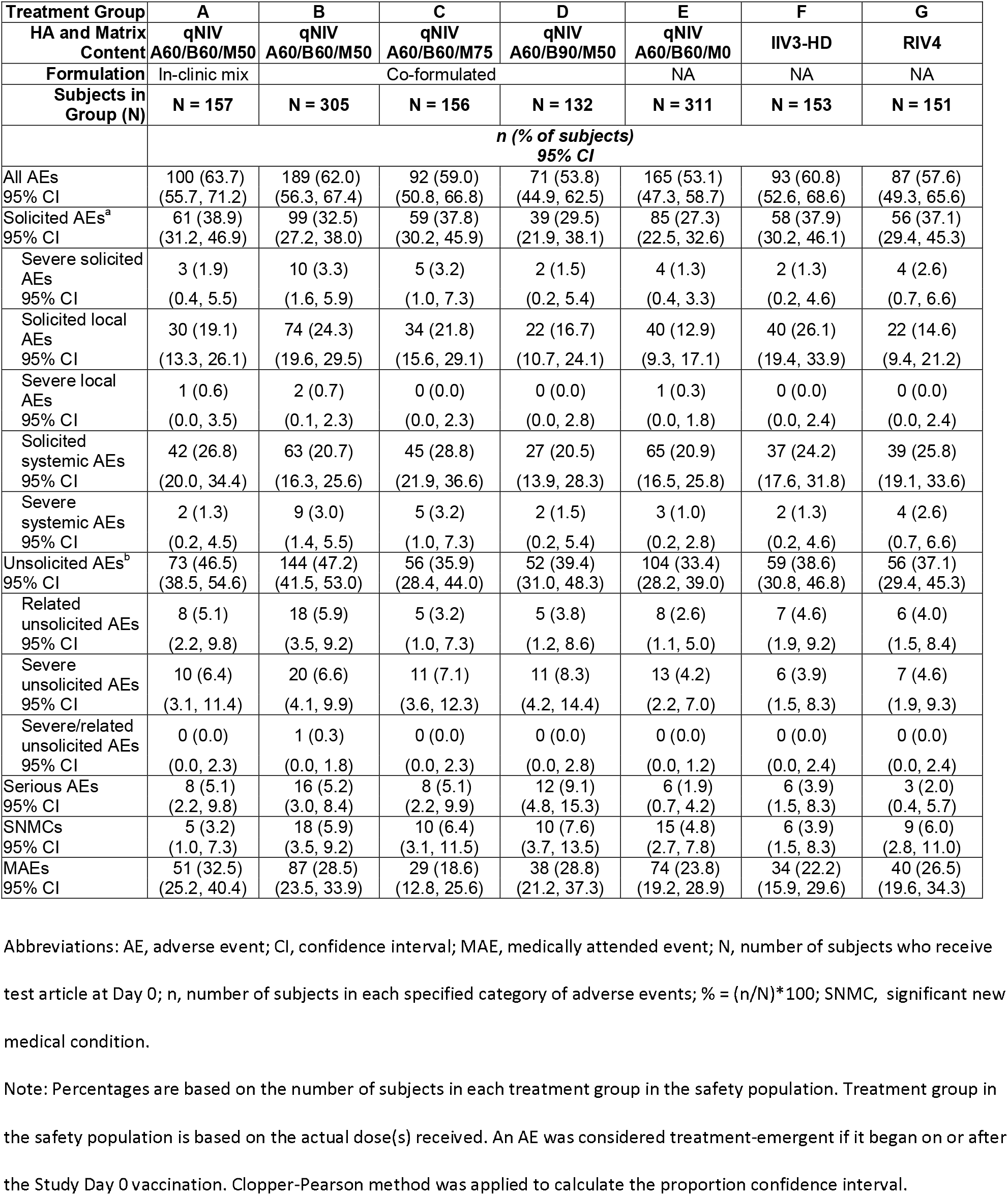
Safety Summary of Adverse Events Post-vaccination Day 0 Through Day 181 – Safety Population.

### Immunogenicity

#### Wild-type hemagglutination inhibition (wt-HAI) antibody responses

The primary objective of demonstrating an adjuvant effect was achieved, with statistically significant increases in wt-HAI antibody responses ranging from 15–29% on 5 of 6 strains evaluated for Group B (qNIV 60 μg HA per strain pre-formulated with 50 μg Matrix-M adjuvant per dose) compared to Group E (qNIV 60 μg HA per strain without adjuvant) (Figure 2).

For all 6 influenza A and B homologous and drifted strains evaluated, Groups A, B, and C (qNIV with 60 Hg HA per strain, mixed in-clinic or pre-formulated, with either 50 μg or 75 μg Matrix-M per dose) showed comparable induction of wt-HAI antibody responses based on Day 28 GMTs, GMRs, SCRs, and SPRs (Table 1; Table S4), indicating that extended pre-formulation of antigen and adjuvant was feasible as it yielded immunogenicity similar to in-clinic mixture immediately before administration. There was no apparent incremental advantage to an increased adjuvant dose of Matrix-M based on wt-HAI antibody responses alone. In contrast, Group D (which received an increased dose of both B antigens, but similar in A antigen content to other groups) compared to other Matrix-M adjuvanted qNIV groups (A, B, and C), did not show improvement in wt-HAI responses against B strains, but instead showed a tendency to reduced wt-HAI antibody response against several A strains, suggesting that an asymmetric increase in the content of B antigens relative to A antigens was not beneficial and potentially interfering.

Based on the comparability of qNIV Groups A, B, and C on wt-HAI responses, Groups B and C were further compared to IIV3-HD and RIV4. Group A was not further considered due to the requirement of an in-clinic mix of antigen and adjuvant. At Day 28, qNIV Groups B and C, compared to IIV3-HD, showed increased wt-HAI antibody responses across a panel of A/H3N2 strains: 40-46% increased against the vaccine-homologous A/H3N2 strain (A/Singapore/INFIMH-16-0019/2016); 18-23% increased against a historically antigenically drifted A/H3N2 strain (A/Switzerland/9715293/2013); and 39-43% increased against a contemporary, antigenically drifted A/H3N2 strain (A/Wisconsin/19/2017) (Figure 3A). Wt-HAI responses to vaccine homologous A/H1N1 and B-Victoria lineage strains were comparable between qNIV Groups B and C versus IIV3-HD. In contrast, across all 6 homologous or drifted strains evaluated, qNIV Groups B and C showed wt-HAI antibody responses comparable to RIV4 (Table 1; Figure 3B). As expected, all treatment groups showed decay of wt-HAI antibody titers at later time points (Day 56, and through Day 182), although between-groups differences were largely preserved (Table S4).

**Figure 3A.**
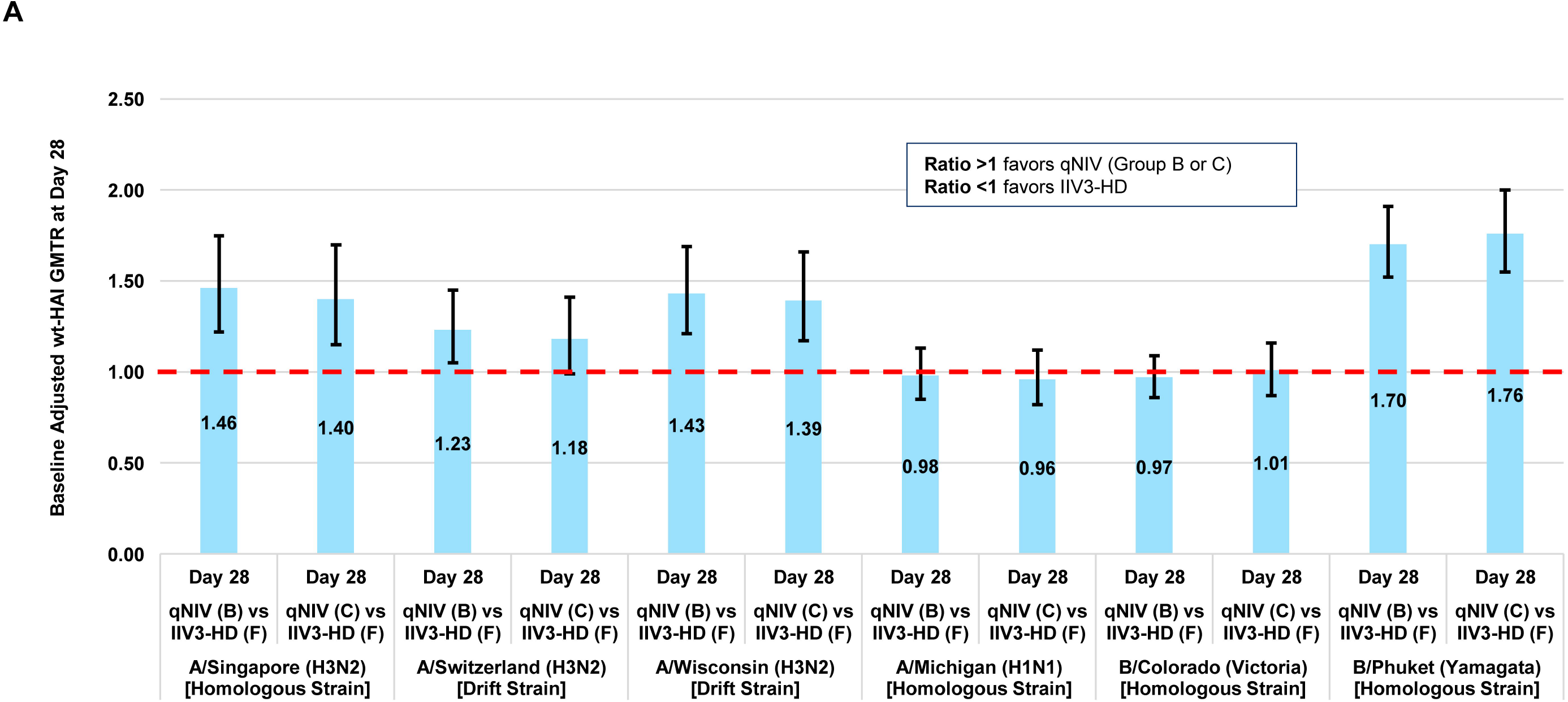
qNIV (Group B or C) compared to IIV3-HD – baseline adjusted ratio of Day 28 wt-HAI GMTs (GMTR) (qNIV [Group B or C]/IIV3-HD [Group F]) Abbreviations: B, Group B; C, Group C; F, Group F; GMT, geometric mean titer; GMTR, ratio of GMTs; IIV3-HD, trivalent highdose inactivated influenza vaccine; qNIV, quadrivalent recombinant nanoparticle influenza vaccine; wt-HAI, wild-type sequenced hemagglutinin inhibition antibody. Full strain names: A/Singapore/INFIMH-16-0019/2016 (H3N2); A/Switzerland/9715293/2013 (H3N2); A/Wisconsin/19/2017 (H3N2); A/Michigan/45/2015 (H1N1); B/Colorado/06/2017 (Victoria Lineage); B/Phuket/3073/2013 (Yamagata Lineage).

**Figure 3B:**
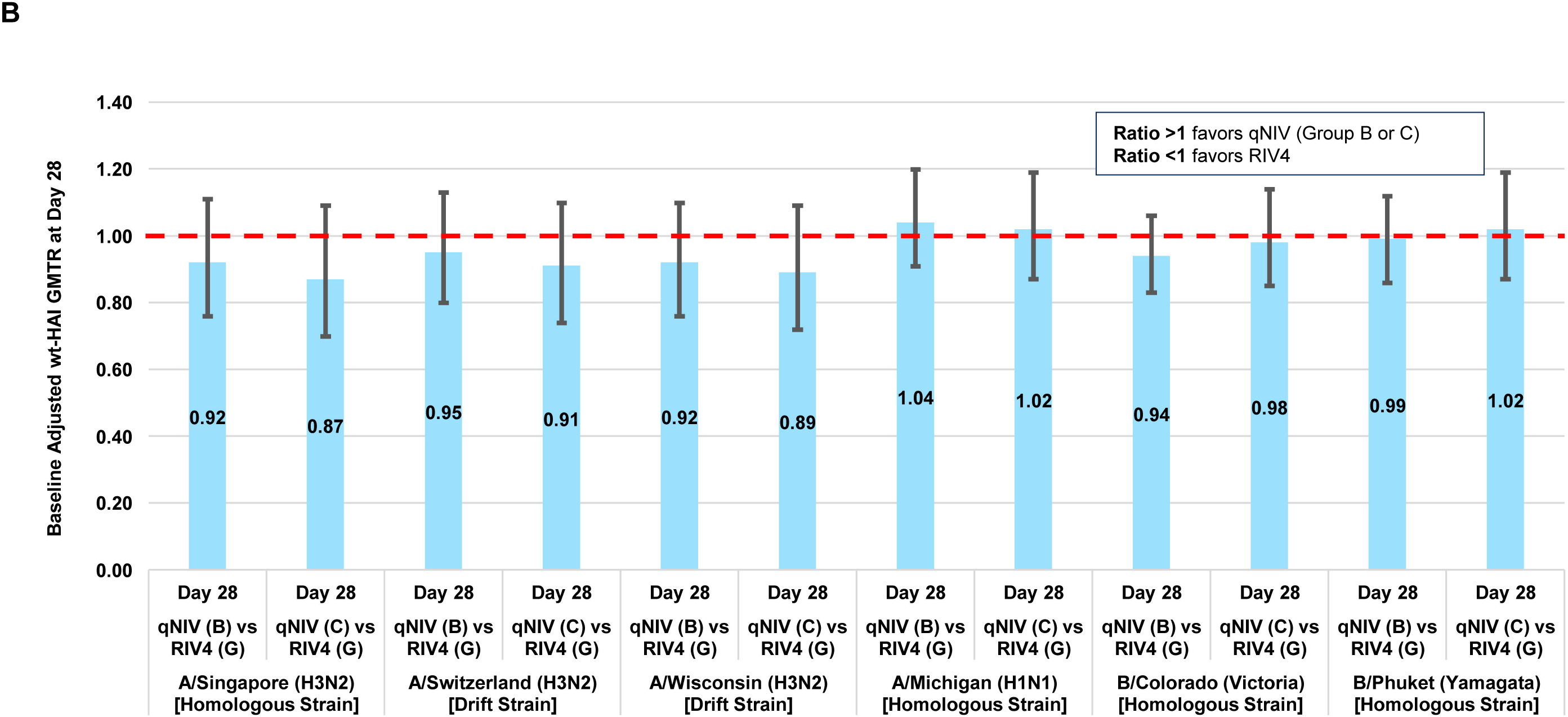
qNIV (Group B or C) compared to RIV4 – baseline adjusted ratio of Day 28 wt-HAI GMTs (GMTR) (qNIV [Group B or C] / RIV4 [Group G]) Abbreviations: B, Group B; C, Group C; G, Group G; GMT, geometric mean titer; GMTR, ratio of GMTs; qNIV, quadrivalent recombinant nanoparticle influenza vaccine; RIV4, quadrivalent recombinant influenza vaccine; wt-HAI, wildtype sequenced hemagglutinin inhibition antibody. Full strain names: A/Singapore/INFIMH-16-0019/2016 (H3N2); A/Switzerland/9715293/2013 (H3N2); A/Wisconsin/19/2017 (H3N2); A/Michigan/45/2015 (H1N1); B/Colorado/06/2017 (Victoria Lineage); B/Phuket/3073/2013 (Yamagata Lineage). *Note: Since Day 56 samples were tested separately from Day 0 and 28 samples, Day 56 titers were adjusted for the long-term assay variability. The adjustment was based on retesting of randomly selected subset 50 subjects of Day 0 samples concurrently with Day 56 samples.

#### Cell-mediated immune (CMI) responses: double- and triple-cytokine producing influenza strain-specific CD4+ T-cells

Pre- and post-vaccination distributions of strain-specific CD4+ T-cell responses for qNIV Group B and C, and IIV3-HD and RIV4, are shown in Figure 4 and described in Tables S5A-C. At Day 7 post-vaccination, polyfunctional phenotypes of double- and triple-cytokine producing influenza vaccine-homologous and drifted A/H3N2 strain-specific responses were induced in all Matrix-M-adjuvant-containing qNIV formulations (Groups A, B, C, D; Table S5A). Among the 4 Matrix-M-adjuvanted qNIV formulations, Group C, with a higher dose of Matrix-M adjuvant (75 μg), showed the greatest induction of post-vaccination double- and triple-cytokine CD4+ T-cell responses across all strains evaluated, and compared to the unadjuvanted formulation of qNIV, demonstrated 11.1-13.6-fold increases (all P<0.01) in Day 7 post-vaccination double-cytokine responses. Remarkably, and uniquely to qNIV Group C, most participants had an influenza strain-specific double cytokine CD4+ T-cell response across all homologous and drifted strains evaluated, indicating a relative absence of CMI “non-responders” (Figure 4A). Compared to IIV3-HD and RIV4, qNIV Group C showed substantially higher post-vaccination fold-rises in inductions of double- and triple-cytokine producing CD4+ T-cells (Table S5A). In terms of between-group differences, qNIV Group C induced 4.1–30.8 and 6.6–31.5-fold higher double-cytokine influenza strain-specific responses at Day 7 post-vaccination compared to IIV3-HD and RIV4 across strains, respectively; and a corresponding 9.9–66.6 and 9.6–14.1-fold higher triple-cytokine influenza HA-specific responses at Day 7 post-vaccination compared to IIV3-HD and RIV4 across strains, respectively (Table S5B and S5C).

**Figure 4A&B:**
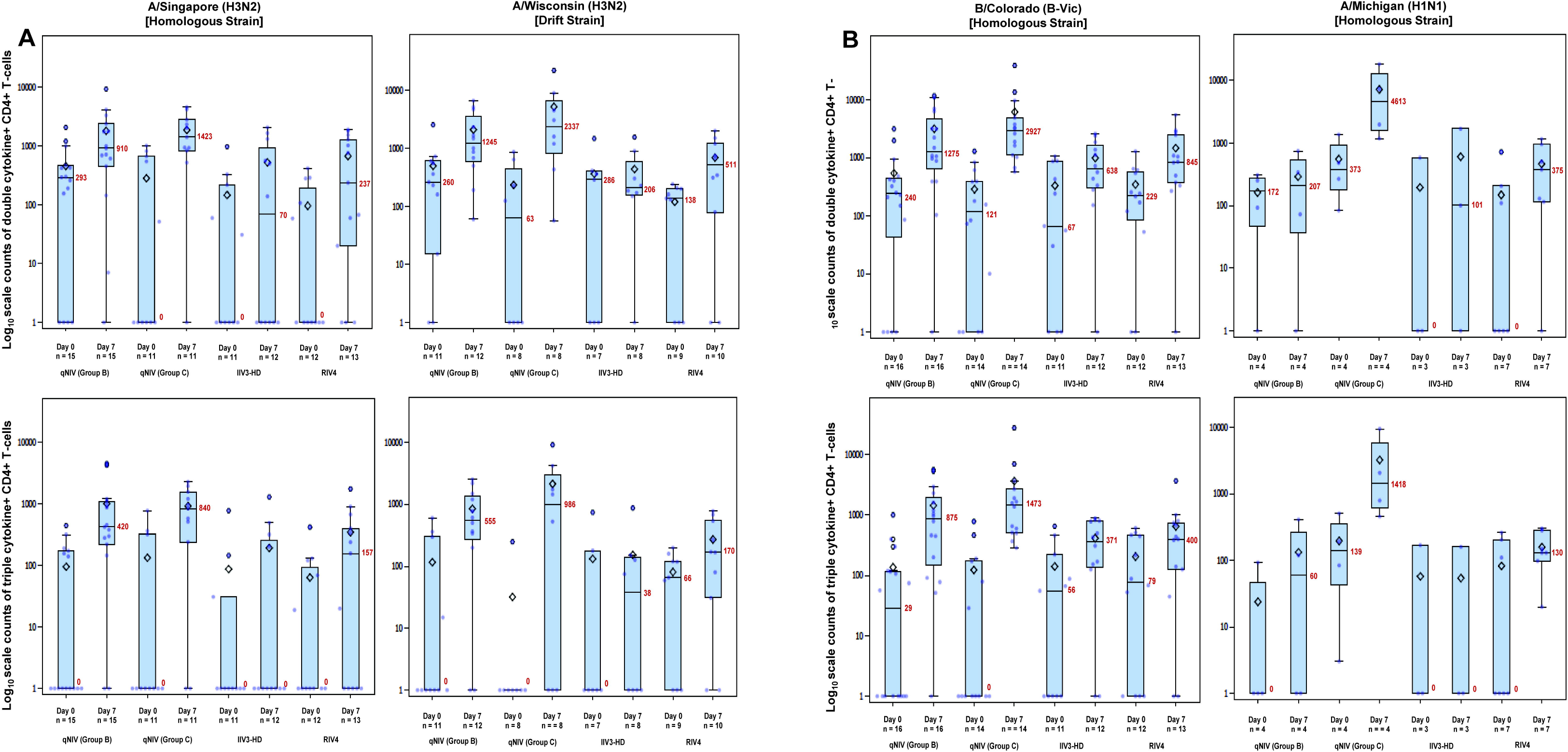
Logio scale counts of double- or triple-cytokine producing strain-specific CD4+ T-cells, by treatment group, time point, and strain. Abbreviations: qNIV, quadrivalent recombinant nanoparticle influenza vaccine; IIV3-HD, trivalent high-dose inactivated influenza vaccine; qNIV, quadrivalent recombinant nanoparticle influenza vaccine; RIV4, quadrivalent recombinant influenza vaccine. Cell-mediated immune (CMI) responses were measured by intracellular cytokine staining (ICCS). Counts of peripheral blood CD4+ T-cells producing interleukin-2 (IL-2), interferon gamma (IFN-γ), and/or tumor necrosis factor alpha (TNF-α) cytokines were measured following in vitro re-stimulation with vaccine-homologous (A/Singapore/FIMH-16-0019/2016[H3N2]; A/Michigan/45/2015[H1N1]; /Colorado/06/2017[Victoria]), or drifted (A/Wisconsin/19/2017[H3N2]) strain-specific recombinant wild-type sequence hemagglutinins (HAs). Panel A shows CMI responses against A/Singapore and A/Wisconsin. Panel B shows CMI responses against B/Colorado and A/Michigan. Boxplots are shown for counts of double-cytokine producing (any 2 of: IFN-γ, TNF-α, or IL-2), or triple-cytokine producing (all 3 of: IFN-γ, TNF-α, and IL-2), strain-specific CD4+ T-cell responses across the 4 strains evaluated using PBMCs obtained from a subgroup of subjects on Day 0 (pre526 vaccination) and Day 7 (post-vaccination). The box plots represent the interquartile range (± 3 standard deviations); the solid horizontal black line represents the median and the number in red indicates the median count of double- or triple-cytokine producing CD4+ T-cells, respectively; and the open diamond represents the mean. Group B is qNIV 60 μg HA x 4 strains with 50 μg Matrix-M1 adjuvant; Group C is qNIV 60 μg HA x 4 strains with 75 μg Matrix-M1 adjuvant; Group F is IIV3-HD; and Group G is RIV4. Note that the number of strains tested for a given participant’s sample was dependent on the number of cells available; thus, not all samples could be tested across all 4 strains.

## DISCUSSION

Seven of the last 9 US influenza seasons have been characterized by dominant or co-dominant circulation of A/H3N2 viruses, with frequent reports of suboptimal field vaccine effectiveness—driven principally by the under-performance of the A(H3N2) component of the vaccine [3,8-11,30]. The burden of A/H3N2-associated morbidity and mortality has been borne disproportionately among older adults [20-22], Existing high vaccine coverage rates in this population suggest that new immunization strategies are urgently required [31]. We have developed qNIV, a novel recombinant HA nanoparticle vaccine, formulated with Matrix-M adjuvant, to address limitations of currently licensed, predominantly egg-derived, seasonal influenza vaccines [26, 28]. We demonstrated in this study that qNIV induced crossreactive humoral immune responses against drifted A/H3N2 viruses in a manner that IIV3-HD did not, and cellular responses against both vaccine-homologous and drifted A/H3N2 viruses in a manner that neither IIV3-HD or RIV4 could. These results suggest that substantial quantitative and qualitative enhancements of the humoral and cellular immune response against seasonal influenza viruses are possible in an older adult population in whom the challenge of a senescent immune system has historically proven difficult for influenza vaccines to overcome, particularly induction of cellular immunity.

A fundamental problem limiting the performance of existing influenza vaccine technologies has been the induction of narrow, vaccine-strain specific immunity [32,33]. This creates vulnerability to classical antigenic drift from a virus adapted to evolve rapidly to evade host immune pressure. The consequences of antigenic mismatch were illustrated by the 2 recent US influenza seasons, which were characterized by the emergence of A/H3N2 drift variants antigenically distinct from the A/H3N2 vaccine strains, and produced estimates of A/H3N2-specific vaccine effectiveness in adults ≥65 years as low as 13% (95% CI:-46–48) and 10% (95% CI:-32–39) during the 2018–2019 and 2017–2018 seasons, respectively [8,9]. Our previous phase 1/2 study of tNIV (conducted in advance of the 2017–2018 US influenza season) demonstrated a 60% improvement in wt-HAI antibody responses induced by tNIV compared to IIV3-HD against the then, newly emerged, antigenically advanced drift variant A/Singapore/INFIMH-16-0019/2016 (H3N2)-like clade 3C.2a1 [28]. In the present study, we showed that against both historic [A/Switzerland/9715293/2013 (H3N2) clade 3c.3a] and contemporary [A/Wisconsin/19/2017 (H3N2) clade 3C.2a2] A/H3N2 drift variants, qNIV demonstrated improved wt-HAI antibody responses relative to IIV3-HD. In contrast, qNIV appeared to induce similarly robust wt-HAI antibody as RIV4 against the same panel of vaccine-homologous and drifted A/H3N2 viruses. Based on previous work with tNIV in ferrets and vaccine-induced bnAbs isolated from mice [27], we posit that breadth of cross-reactivity induced against drifted influenza strains may, in part, be mediated by the induction of bnAbs that interact with conserved HA head epitopes, both near and distant to the receptor binding domain, as well as conserved HA stem epitopes [26,27].

A second critical challenge limiting vaccine performance is the increasingly recognized problem of egg-adaptive antigenic changes arising from traditional egg-based manufacturing methods [16,18]. While not a new challenge, the consequences of this problem have gained focus with data characterizing specific egg-adapted HA antigenic site mutations as having deleterious effects on A(H3N2) vaccine immunogenicity and effectiveness [6, 12-15]. The potential adverse impact of egg-adaptive mutations on vaccine immunogenicity, and the capacity of qNIV to overcome this problem by preserving wt HA sequences, was illustrated in the phase 1/2 study by the substantially enhanced neutralizing antibody responses induced by tNIV relative to IIV3-HD against *wt* sequenced A/Singapore/INFIMH-16-0019/2016 (H3N2) virus; whereas, when neutralizing antibody responses were assessed against *egg-adapted* A/Singapore/INFIMH-16-0019/2016 (H3N2) virus, both vaccines appeared to perform comparably (Figure S1). These data highlight not only the problems of egg-derived influenza vaccines, but also the corresponding problem of using egg-derived viral reagents in either HAI or neutralization assays, which may lead to a biased assessment of vaccine immunogenicity in favor of egg-derived vaccines, and away from wt virus relevant immune responses that may better predict clinical protection against circulating viruses [34].

A third critical challenge—of particular importance for older adult immunization—has been the limited induction of cellular immunity by currently licensed influenza vaccines. A randomized clinical trial (RCT) from Canada in older adults evaluated CMI responses to 4 licensed inactivated seasonal influenza vaccines—standard subunit, MF-59 adjuvanted subunit, standard split virus, or intradermal split virus— and found that all 4 vaccines had similar but limited induction of CMI responses, including no meaningful post-vaccination increases in CD4+ and CD8+T-cells expressing IFN-γ, IL-2, or IL-10 [24], Similarly, a recent RCT from Hong Kong in older adults comparing CMI responses of 3 “enhanced” influenza vaccines (IIV3-HD, RIV4, and MF-59-adjuvanted IIV3) relative to a standard dose IIV3, also found only modest Day 7 post-vaccination induction of strain-specific IFN-γ+CD4+T-cell responses (range 1.0-2.6 fold-increases for enhanced vaccines; and 1.2-1.8 fold-increases for standard dose IIV) [25]. The reported magnitude of these post-vaccination CD4+ T-cell responses were in line with similar observations regarding IIV3-HD and RIV4 in our study. These findings come amid a growing recognition that CD4+ and CD8+ T-cell responses are important for modulating influenza disease severity and conferring a breadth of vaccine protection [35-37]. Failure to induce potent cellular immunity to influenza is a problem of increased consequence in older adults because age-related declines in T-cell function and concomitant age-related increases in frailty may converge to both diminish vaccine response and increase the risk of serious complications of influenza virus infection [38-40]. Notwithstanding the limited sample size for CMI assessments in this study, we observed statistically significant activation of influenza HA-specific polyfunctional CD4+ T-cell responses by qNIV in an older adult population, which could be restimulated by either vaccine-homologous or drifted HA antigens, in a manner not previously reported for existing seasonal influenza vaccines.

In conclusion, this study showed that Matrix-M-adjuvanted recombinant qNIV was well tolerated and could markedly enhance *both* broadly cross-reactive antibody *and* cellular immune responses. This approach is being further studied in a phase 3 trial, and has been granted access to the accelerated approval pathway by the FDA.

## Data Availability

The datasets generated during and/or analyzed during the current study are not publicly available as analyses are ongoing and will be used for regulatory submissions for the vaccine described; but may become available in the future from the corresponding author on reasonable request.

## FUNDING

This work was supported by the sponsor, Novavax Inc. (Gaithersburg, MD, USA).

## ACKNOWLEDGMENTS

We thank the study participants, the investigators from the clinical trial sites, members of the sponsor’s and contract research organization (CRO) team, and the Clinical Immunology laboratory for their contributions to the trial. Editorial assistance on the preparation of this manuscript was provided by Phase Five Communications, supported by Novavax, Inc.

## POTENTIAL CONFLICT OF INTEREST

Vivek Shinde, Joyce Plested, Iksung Cho, Jamie Fiske, Mingzhu Zhu, Shane Cloney-Clark, Haixia Zhou, Bin Zhou, Nita Patel, Michael J Massare, Gale Smith, Louis Fries, and Gregory M Glenn are current employees of Novavax, Inc. Amy Fix, David Nigel Thomas, Michelle Spindler, Rongman Cai, Nan Wang, and Xuan Pham are former employees of Novavax, Inc.

## Clinical Trials Registration

ClinicalTrials.gov NCT03658629.

## Notes

### Clinical Trial

NCT03658629

